# Risk Assessment and Mapping in a Space Analog Station: Collaborations to Ensure Safety and Minimize Failures

**DOI:** 10.1101/2024.11.29.24318161

**Authors:** Iris Campanella Cabral, Ana Carolina Seixas de Oliveira Santana, Júlio Francisco Dantas de Rezende, Soraya Chucair, Gabriel Matheus Dutra Santos, Cristina Ribas Fürstenau

## Abstract

The development of Map Assessments, especially in analog space missions, plays a fundamental role in identifying, assessing, and mitigating potential threats and challenges that may arise during space operations in extraterrestrial environments. These missions are designed to simulate real space flight conditions, allowing space agencies and exploration teams to test and refine procedures, technologies, and protocols before implementing them in actual space missions. During the risk analysis, various factors are considered, including: i) Environmental Risks: the context in which the mission will occur, as extreme weather conditions; ii) Health and Safety Risks: health and safety of the team members, as exposure to radiation; iii) Operational Risks: associated with operational procedures, communications, resource management, team coordination, and decision-making during the mission; iv) Psychosocial Risks: interpersonal conflicts, and emotion management along with the crew. Based on the analysis of these risks, strategies and mitigation measures are developed to minimize the likelihood of adverse events and their potential consequences. This study aims to analyze occupation risks, divided into physical, chemical, ergonomic, biological, and mechanical, at the analog space station Habitat Marte, located in Rio Grande do Norte, Brazil. As a result, various sources of danger and vulnerabilities in analog space operations were identified, which include: a. identification of the biological risk group, which includes symptoms associated with transmitted diseases or infections, the possibilities of contamination and spread of transmissible diseases; b. identification of the chemical risk group, which includes the presence of harmful gases, such as carbon dioxide and methane, symptoms of exposure, such as breathing difficulties; c. Identification of the mechanical risk group, where insect bites, possible fires, structural collapses, d. Identification of the physical risk group, which includes exposure to extreme temperatures, dehydration, fatigue and falls, e. Identification of the ergonomic risk group, strongly present in issues such as work in confined spaces and repetitive movements, f. Identification of the psychological risk group, which ultimately encompasses stress due to confinement, lack of rest, and fear of incidents during the journey. By conducting this comprehensive and multidisciplinary risk analysis, space agencies and exploration teams can significantly increase the safety, efficacy, and success of their future missions in space. These results provide a solid foundation for the continuous development of risk management strategies and safety measures for long-duration space missions, contributing to the viability and success of future human expeditions to Mars and beyond.

## 1. Introduction

Throughout life, human beings are exposed to various types of hazards with different levels of associated risk, often without full awareness or recognition of their presence. These hazards can be present in domestic environments, public spaces, and especially in the workplace [1]. Occupational risks are defined as adverse events or conditions in the work environment that arise from both individual actions and organizational factors. These risks can compromise human health, leading to injuries, illnesses, or even fatalities [2].

Space exploration has been the subject of studies that aim to reach new heights in scientific and technological advancement. The pursuit of human exploration beyond Earth has entered a crucial phase with NASA’s Human Spaceflight Plans, detailed in the Campaign Report, which projects potential orbital missions to Mars by the end of the 2020s [3]. However, significant challenges remain, including ensuring the physical and mental well-being of the crew. [4]. This encompasses managing stress, preventing cognitive problems, mitigating sensory deprivation, and ensuring effective communication and team dynamics [5].

Analog stations provide dedicated astronaut training environments, replicating some complexities faced in manned space missions [6]. These analog missions are often conducted in environments that mimic target characteristics, such as temperature, isolation, topology, and mineralogy [7]. They offer invaluable opportunities for researchers, scientists and explorers from different fields to develop critical skills such as problem solving under pressure, efficient decision making and effective teamwork under conditions of isolation and confinement [8,9]. The risks present at these stations help train and select astronauts capable of handling extreme challenges during missions to Mars and interplanetary journeys [10]. However, the controlled presence of risks in these environments is essential so that training can realistically simulate the conditions that will be faced in space missions. In this sense, the identification of these risks in environments similar to Mars, as well as the development of strategies to minimize them, is essential to ensure both the safety of the crew and the success of the missions.

This study focuses on the Habitat Marte analogue station, located in the rural area of Caiçara do Rio do Vento, Rio Grande do Norte, Brazil. Designed as an immersive training complex, Habitat Marte simulates manned missions to Mars and serves as an interdisciplinary research center with applications for Martian base development and aerospace advancement. The station offers infrastructure for long-term stays, leveraging local biome characteristics to simulate Martian conditions, including soil, climate, and atmosphere studies, simulated spacesuit expeditions, and extravehicular activities. A key challenge is achieving self-sustainability through aquaponics for food production, continuous water supply and reuse, and sustainable energy generation.

Therefore, this study aims to investigate occupational risks at the Habitat Marte analogue station, identify implemented safety mechanisms, and map risks associated with extravehicular, intravehicular, and underwater extravehicular activities conducted in the complex.

## 2. Theory

In the literature review, it was crucial to highlight studies addressing occupational risks, with a specific focus at the end on psychological risks, as well as research related to the development of risk maps in the aerospace sector.

### 2.1 Occupational Risks

Occupational risks are assessed based on the frequency, probability and severity of the consequences of adverse events caused by risk factors. These factors, alone or in combination, can cause immediate or future damage, and may affect the physical and mental health, safety and well-being of the people involved [11]. Risks represent exposure to danger, that is, the effect of interaction with sources of damage. Good management of exposure to these hazards can mitigate or minimize risks [12]. There is still no clear evidence in the literature about specific occupational risks in Mars-like stations. However, it is known that space travel involves several risk factors that, depending on their intensity and duration, can cause chronic damage to the crew’s health [13,14]. Patel (2020) highlights with the main risks are space radiation, which can result in cancer, cardiovascular disease and cognitive decline; Space Flight Associated Neuro-ocular Syndrome (SANS), which causes cardiovascular, cerebral and oculomotor changes; in addition to orthostatic intolerance upon returning to Earth [15]. Risks to behavioral health, performance, inadequate nutrition, sarcopenia, osteopenia and an increased risk of falls and fractures during extravehicular activities are also observed [16]. Studies indicate that, due to the extreme conditions and prolonged exposure time, the human body is not yet completely prepared for a manned mission to Mars. This demands the development of new technologies to mitigate short- and long-term impacts [17].

The way people, objects or systems are exposed to danger is decisive for mitigating the associated risk. Factors such as exposure time and nature of the risk must be taken into account, in addition to the routes of penetration, which can worsen the aggressiveness of the risk. Occupational risks are classified according to their nature, concentration, intensity and time of exposure, as: i) physical, ii) biological, iii) mechanical, iv) chemical and v) ergonomic [12]. According to the NR-9 Risk Prevention Program [18], physical risks are the various forms of energy that may be exposed to workers, such as: noise, radiant heat, humidity, cold, abnormal pressures, ionizing radiation and non-ionizing, vibrations, as well as infrasound and ultrasound. Chemical risks refer to substances that can enter the body through breathing or through contact with the skin and ingestion, such as dust, smoke, gases and vapors. Biological risks involve microorganisms, such as bacteria, fungi, parasites, protozoa and viruses, which can cause illnesses when they come into contact with workers. Mechanical risks are associated with inadequate conditions in the workplace, such as improper physical arrangement, lack of protection on machines, defective tools and poor lighting, which can result in serious accidents and physical problems. Ergonomic risks arise from the interaction between the worker and their environment, including inadequate postures, excessive physical effort and intense work rhythms, which can lead to fatigue, muscle pain, back problems and diseases such as hypertension, diabetes and RSI/WMSD.

Ergonomic risks also encompass psychosocial risks, which, despite having often been neglected in several areas, including space exploration, are now recognized as essential to ensuring well-being and safety in the workplace [19]. Conditions such as prolonged stress, isolation, excessive control over work and monotony can contribute to the emergence of psychological disorders such as depression, anxiety, apathy and cognitive decline, especially in extreme situations such as long-duration space missions [5]. Mental health care is crucial to mitigate these risks, and psychological support strategies, such as the development of new technologies and self-assessment tools, are increasingly necessary to preserve crew members’ emotional balance [5,20]. In this context, analogue stations play a crucial role in providing a controlled environment to study and mitigate these psychological risks, enabling the development of countermeasures and psychological support strategies that are essential for long-duration interplanetary missions [21,22]. These simulations help test methods and systems for managing stress and loneliness, which are fundamental to ensuring the well-being of astronauts.

Despite the complexity of identifying all hazards, risk identification techniques have advanced over the decades, considering each specific process and area, which improves the effectiveness of assessments [23]. Traditional risk calculation methods remain largely based on simplified formulas, such as risk = probability × severity, or risk = exposure × probability × consequence [24]. These formulas are used in risk assessment software used by public and private companies. Based on these indices, it is possible to develop risk matrices and maps, which are essential for occupational risk and safety management [24].

### 2.2 Risk mapping

In the 1960s, Italy had high rates of workplace accidents, which led unions to come together with the aim of solving the problem [24]. Thus, in the 1970s, the Environmental Risk Map emerged in Europe, which spread throughout the world and arrived in Brazil in the 1980s, with the main objective of gathering essential information to establish a diagnosis of the safety and health situation at work. within companies [25]. The Risk Map is a graphic representation that highlights various factors present in workplaces, which can cause damage to workers’ health, being an essential tool for communicating in a simple and organized way the results of complex risk assessments to workers, the community location and crew.

Regulatory Standard 05, which deals with CIPA, regulates the Risk Map in Brazil. Although the requirement has changed with recent updates, the Risk Map continues to be the most used visual resource to represent the risks present in the workplace [18]. It aims to help reduce the occurrence of accidents and provide an easy visualization of the risks existing in the organization [26].

It is relevant to discuss the specific applications of risk mapping in the space area, especially in missions involving extreme and complex environments. The risk mapping technique plays a fundamental role in identifying and mitigating the dangers inherent to space missions. A notable example of this approach is the Probabilistic Risk Assessment Procedures Guide for NASA Managers and Practitioners [27]. This guide provides a comprehensive methodology for probabilistic risk assessment (PRA), which is widely used by NASA to ensure the safety and success of space missions. PRA integrates detailed analyzes of the potential risks associated with each component of a mission, including technological systems, human factors and the space environment, enabling the anticipation and mitigation of failures that could compromise the mission.

Although mapping risks associated with work on analogous space stations is still an emerging area, it already benefits from consolidated approaches and concepts in related areas, such as safety and risk management in engineering projects.

## 3. Material and methods

The data collected in this study were categorized into three groups: (a) Direct observations made by the first author, who was a crew member of the Gaia Mission conducted from November 11 to December 12, 2022; (b) Detailed interviews with the Analog Station manager; (c) Collection of geospatial data on extravehicular activities using GIS software.

### 3.1 Brief description of the study area

The study focuses on the Habitat Marte analogue station, located in the city of Caiçara do Rio do Vento (5º08’06” S, 35º59’06” W), in the state of Rio Grande do Norte, Brazil (Figure 2). This state is situated in the eastern part of the Northeast region of Brazil, covering an area of 52,811.126 km^²^, divided into 167 municipalities, and bordering the states of Ceará and Paraíba [28]. The region is part of the Caatinga biome and has a semi-arid climate, characterized by high temperatures, a low annual temperature range, and minimal rainfall, with long periods of drought [29].

**Fig. 1.**
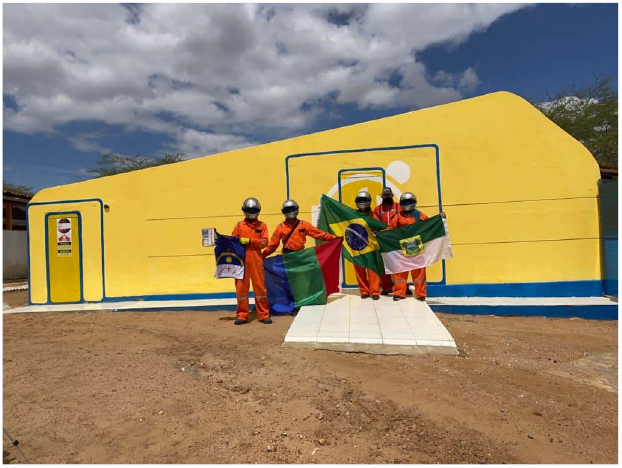
Front view of the Mars analogue research station Habitat Marte, located in the Brazilian semi-arid region.

**Fig. 2.**
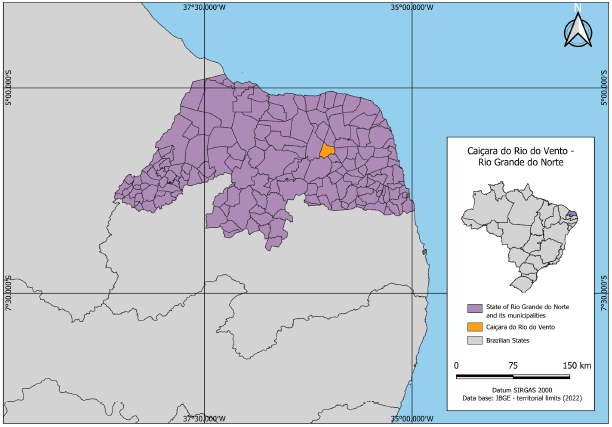
Location map of the State of Rio Grande do Norte. Highlighted in orange is the municipality of Caiçara do Rio do Vento. Source: Author’s own elaboration.

### 3.2 Research Preparation Steps

To understand the current scenario of risk management at the Habitat Marte analogue station, it was essential to review the work processes and activities related to the station’s operations. The methodology employed includes both descriptive and exploratory studies. The descriptive study observes, records, and correlates facts or phenomena without manipulation. The exploratory study provides detailed descriptions of the situation and the relationships between its elements, focusing on defining objectives and gathering further information on the topic of study [30].

Therefore, the author of this study conducted a detailed analysis of the location, observing the characteristics of all internal and external compartments as well as the EVAs where the missions are carried out. Observations were made regarding individual and collective safety procedures used by the crew, along with photographic documentation of the site. Additionally, a detailed interview was conducted with the manager of the Habitat Marte Analog Station, Prof. Dr. Júlio Rezende, to gather local information on the following points: (a) The number of people involved in the missions; (b) The duration and description of each activity; (c) The presence of risk identification and control tools; (d) The existence of prior training; (e) Evidence and records of accidents; (f) The availability of emergency supplies at the station; (g) The number of health professionals involved in the missions.

#### 3.2.1 Risk analysis at IVA and UEVA

To differentiate the occupational risks associated with the activities at the space analog station, the risk classification table provided by NR-9 [18] was used. This table categorizes risks by color and severity. The analysis considered physical risks (such as noise, heat, humidity, and radiation), chemical risks (such as dust, fumes, gases, vapors, and mists), biological risks (such as fungi, viruses, bacteria, protozoa, and insects), ergonomic risks (such as repetitive movements, excessive pace, physical strain, and poor posture), and mechanical risks (such as inadequate lighting, fire hazards, and electrical risks). After classification, the magnitudes of these risks were assessed as small, medium, or large, as shown in Table 1.

**Table 1.**
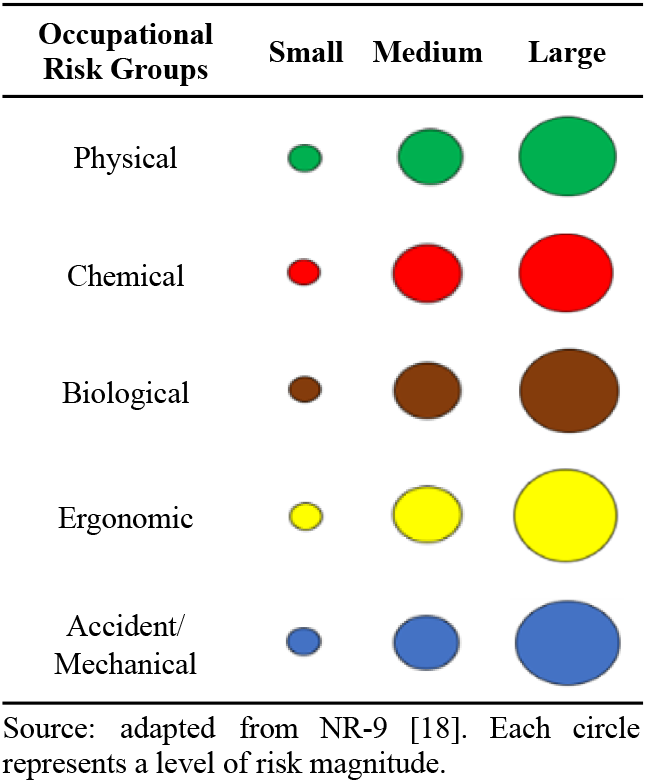
Groups of occupational risks and their magnitudes.

When preparing a risk map, it is essential to consider the different forms of exposure to hazards, such as direct contact, indirect contact, percutaneous contact, airborne contact, or ingestion [1]. Each of these forms of exposure has varying probabilities of occurrence; for example, ingestion is much rarer than direct or airborne contact. Each type of exposure will have an area of influence that must be considered when creating the map. Therefore, by assigning a severity score (S) from 1 to 4 and a probability (P) from 1 to 4, within a defined area based on the space needed for a particular type of contact to occur, the final risk was calculated using the simple matrix method demonstrated below:

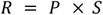

Where P and S are defined according to Table 2:

**Table 2.**
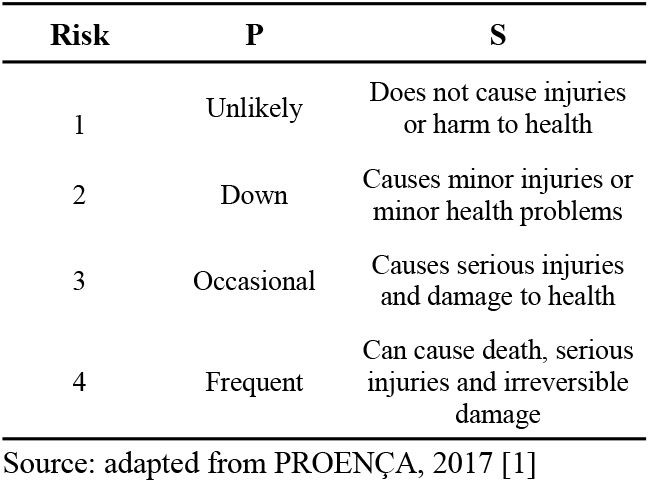
Probability and Severity Values according to the Simple Matrix method.

From the combination of probability categories with severity categories, the Risk Dimensioning Matrix is derived, as shown in Table 3, providing a quantitative indication of the level of risk for each scenario proposed in the analysis of occupational risks on the risk map.

**Table 3.** Risk Dimensioning Matrix.

| P | G |  |  |  |
|---|---|---|---|---|
|  | 1 | 2 | 3 | 4 |
| 1 | 1 | 1 | 2 | 2 |
| 2 | 1 | 2 | 2 | 3 |
| 3 | 2 | 2 | 3 | 3 |
| 4 | 2 | 3 | 3 | 3 |

Source: adapted from PROENÇA, 2017 [1]

After determining the magnitude of the risk, it is necessary to classify it to assess whether it is acceptable, acceptable with need for improvement and attention, or unacceptable. In this classification, risk level 1 corresponds to a small magnitude and is acceptable, though still requiring improvements. Risk level 2 has a medium magnitude, being acceptable but requiring improvements and attention. Risk level 3 represents a high magnitude, making it the most critical and unacceptable. With the combination of these criteria, it is possible to determine the acceptability and measure the magnitude of each risk, regardless of its category.

Based on this assessment, a detailed floor plan of all facilities at the analogue station was developed using the online software Floorplanner (https://floorplanner.com/pt). Based on the classification shown in Table 3, a risk map was created with the objective of facilitating the rapid identification of environmental risks at the station. This map aims to raise awareness and inform crew members, workers, and those responsible for preparing and implementing training.

#### 3.2.2 Risk analysis in EVAs

The paths of extravehicular activities (EVAs) were evaluated in detail at four specific destinations: Lava Cave Habitat (coordinates [−5.80303, −36.01043]), Pico do Cabugi (starting point at [−5.69058, −36.31982] and destination at [−5.70359, −36.32231]), Lago Ceres ([−5.80423, −36.01043]), and Delta Cave ([−5.80482, −36.01173]). The classification of risks associated with EVAs followed the criteria established by NR-9, as detailed in section 3.2.1, with the addition of specific symbols for each type of risk, as described in Table 4. This modification was made to improve clarity in the representation of the various risks identified during the EVAs, as proposed by this research.

**Table 4.**
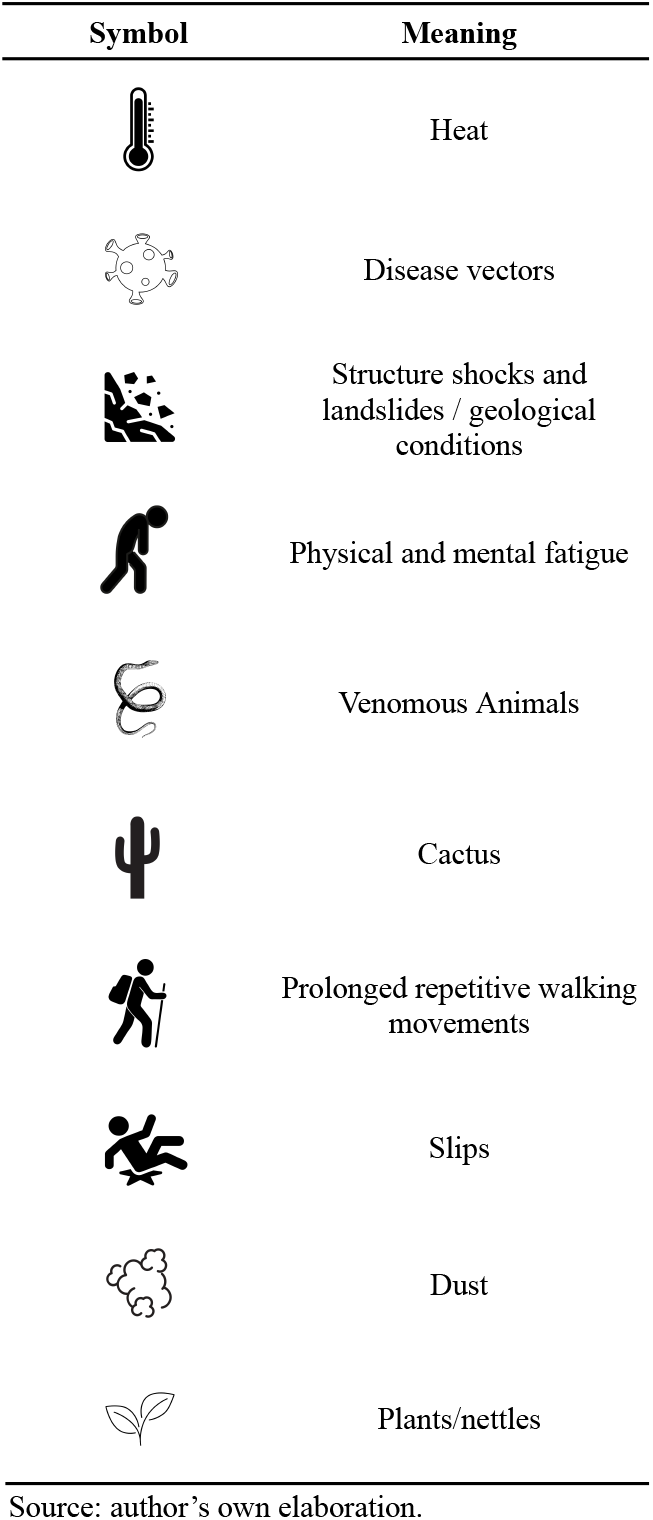
Meaning of symbols used to represent risks.

| Symbol | Meaning |
| --- | --- |
|  | Heat |
|  | Disease vectors |
|  | Structure shocks and landslides / geological conditions |
|  | Physical and mental fatigue |
|  | Venomous Animals |
|  | Cactus |
|  | Prolonged repetitive walking movements |
|  | Slips |
|  | Dust |
|  | Plants/nettles |

Source: author's own elaboration.

As it was not possible to prepare a floor plan for the EVAs, maps were generated and adapted using QGIS software (version 3.34.7). In addition to the observations made during the mission, which helped assess risks, this approach allowed for a detailed geotopographic analysis of the locations, generating data on slope, altitude, terrain characteristics, relief, and distances to key points. The digital terrain model, in GeoTiff format, was obtained through SRTM and accessed via INPE’s

Topodata project (http://www.dsr.inpe.br/topodata/). Additionally, DEMs were provided by IBGE (https://www.ibge.gov.br/geociencias/downloads-geociencias.html) and the MapBiomas project (https://brasil.mapbiomas.org/downloads/).

## 4. Results and Discussion

The results presented in this section refer to the risk mapping of IVAs, UEVAs, and EVAs at the Habitat Marte analogue station.

### 4.1 Intravehicular Activities

#### 4.1.1 Description of activities carried out

The station’s long-duration missions vary in length, with breaks for meals and rest typically occurring between 11 p.m. and 6 a.m. Cleaning and maintenance activities are carried out before, during and after space simulation missions, general cleaning and maintenance of the station is carried out. The station operates on an infrastructure of approximately 460 m^²^ The structure has side windows that favor predominant natural lighting. The station has been in operation for 6 years (since December 2017) and one of the main characteristics of this station is the diversity of tasks developed by the same employee, which involve: i) research, ii) data monitoring, iii) equipment maintenance and iv) general administration of the station. The team typically hosts around a group of 2 to 13 people each month, varying depending on demand. The station has two doctors on duty for 24-hour remote care, offering support in emergencies and health monitoring via telemedicine.

##### 4.1.1.1 Greenhouse (BioHabitat)

This sector is dedicated to growing plants in aquaponics systems (Fig. 3), which integrate plant cultivation and fish farming in a symbiotic system. The plants grown on site are intended for personal consumption, with a predominance of species such as chives, lettuce, basil, sweet potatoes and chickpeas. The greenhouse used for this cultivation has approximate dimensions of 28 x 10 meters, with its walls built with recycled tiles and polycarbonate sheets. Currently, the greenhouse is partially uncovered, presenting a 3×10m covered area. During September 2024 was under development of entire full coverage, ensuring complete protection of the growing season

**Fig. 3.**
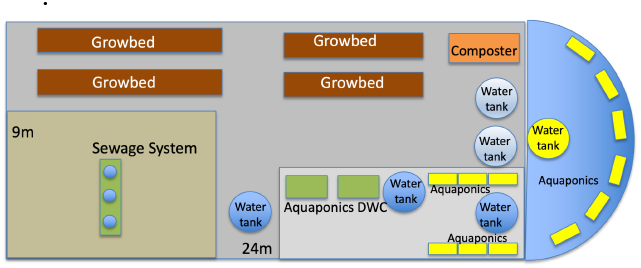
Habitat Marte greenhouse, with aquaponics system.

##### 4.1.1.2 Bedrooms

Dormitories are spaces intended for crew members to rest and recover. There are 3 bedrooms for rest, with bedroom 1 equipped with a 1 bunk bed and 2 beds, bedroom 2 equipped with a bunk bed and bedroom 3 with 3 bunk beds and 1 bed. Activities include regular cleaning, maintenance of the environment, and ensuring a controlled climate with air conditioning for the comfort of crew members. The walls of the bedrooms are made with plastic sheets made from reused material from toothpaste tubes. During the mission, the presence of arachnids, specifically scorpions and spiders, was noted in these compartments.

##### 4.1.1.3 Kitchen/ dining/ meeting room

The kitchen is the sector where meals are prepared for the entire crew, meals take place during the morning, afternoon and night. Prepared meals are easy-to-prepare foods that aim to simulate meals at real stations, such as instant noodles and astronaut paste. The kitchen is equipped with a refrigerator, microwave, sink, cabinets for food storage, table, chairs for social interaction between participants and a type C fire extinguisher.

##### 4.1.1.4 Bathrooms

There is one bathroom at the station, equipped with individual lockers, a sink, a mirror, toilets, a water shower, and a toilet paper holder. During the mission, traces of small insects such as mosquitoes, beetles, and arachnids were noticed, coming from the external area and attracted by the light..

##### 4.1.1.5 Decompression

The decompression room is used to simulate the crew’s transition between the external and internal environment at the station, although there is no real decompression system.

##### 4.1.1.6 Marslab Office

The Marslab workshop is a dry laboratory used for local soil studies, as well as serving as a storage space for a great diversity of tools, such as hammers and wrenches. The laboratory also houses construction materials and maintenance support, such as screws, nuts and paints. Furthermore, Marslab is a collection and analysis point for soil and rock samples, as well as recycled materials, which are studied to evaluate their application in construction projects.

#### 4.1.2 Identification of critical points

##### 4.1.2.1 Biological risk

Within the Habitat Marte analogue station, the possibility of spreading contagious diseases and infections among crew members is a significant concern, given the prolonged isolation and constant coexistence in closed environments [31]. To minimize this risk, it is essential to collect detailed medical records and check the vaccination status of all crew members at least one week before the start of the mission. This preparation makes it possible to identify and mitigate biological risks even before participants arrive at the station. Although on a real mission to Mars, the crew would always be in constant proximity, advance knowledge about each crew member’s health is crucial to reducing the likelihood of disease outbreaks during the mission. Frequent hand hygiene and adequate ventilation of internal compartments complement these measures, helping to maintain the health of the crew during the mission.

The shared use of toilets at the station is another critical point. Brazilian Regulatory Standard 24 [32] requires the separation of bathrooms by gender, which helps to minimize risks. Furthermore, it is crucial to regularly clean and disinfect these areas, using products with disinfectant action, especially in high-traffic areas such as bathrooms and kitchens. Cleaning with detergent, although effective in removing dirt, may not be sufficient to eliminate pathogens, especially considering the large number of people who frequent the station. To ensure adequate disinfection and minimize the risk of contamination, it is recommended to use cleaning products with disinfectant action, especially in high-traffic areas, such as bathrooms, kitchens and common areas.

Additionally, in the greenhouse section, which houses an aquaponics system, there is a high potential for biological risk. Although it is an innovative and sustainable solution for in-season food production, this system can be a potential biological risk due to the presence of fish and plant species. The proximity of the species and the management of the system need to be carefully monitored. Any new fish introduced into the system must go through a quarantine period to ensure that they are not carrying diseases that could spread to other fish or even the greenhouse environment. Furthermore, fish deaths require strict tank removal and disinfection procedures to prevent the proliferation of pathogens. Searches for Mori (2019) show that fish and plant pathogens can spread through systems aquaponics and that physical disinfection methods such as ultraviolet (UV) irradiation, media filtration, membrane filtration and blue light-emitting diodes can help control transmission [33].

Therefore, regular cleaning of the greenhouse and aquaponics system, as well as maintenance and monitoring of filtration systems are crucial to maintaining a healthy environment. Furthermore, it must be ensured that no chemicals are used on the soil or water to avoid risks to both fish and plants. The biological risks present in the station’s common areas can be transmitted via the respiratory, cutaneous or digestive routes, with airborne transmission being the most worrying. These risks were classified as level 2, considering their probability, which may be occasional due to the absence of health questionnaires before missions, and their severity, which may result in injuries or mild health disorders, implying a moderate risk that can be managed with adequate PPE and rigorous disinfection and decontamination procedures.

It is crucial to develop and implement specific protocols to minimize the risk of disease transmission and contact with potential vectors of pathogenic microorganisms. These protocols must include regular training before missions, simulations of contamination scenarios, and the creation of rapid emergency response mechanisms, considering the significant distance between Habitat Marte and medical facilities. In the context of a space analog mission, it is crucial to distinguish between biological hazards that must be eliminated completely and those that, although minimized, must be maintained as part of training. Risks such as the lack of adequate disinfection in common areas and the improper handling of organic waste, such as the inadequate removal of dead fish, must be eliminated, as they do not contribute to training and can compromise the health of crew members. On the other hand, controlled exposure to respiratory diseases and the management of the aquaponics system are important to simulate living in constant proximity and the management of sustainable resources, preparing the crew for similar situations on Mars.

Finally, the station’s equipped pharmacy, containing essential medications such as anti-inflammatories and anti-allergy medications, serves as additional support for the management of any health complications that may arise, reinforcing the safety and well-being of the crew during the mission.

##### 4.1.2.2 Chemical risk

Air conditioning systems in Habitat Marte station dormitories may pose chemical hazards due to the use of refrigerants such as hydrofluorocarbons (HFCs). Although HFCs have been adopted as an environmentally safer alternative to chlorofluorocarbons (CFCs) [34], acute exposure to these compounds, especially in cases of leaks, can cause symptoms such as dizziness, headaches and, in more serious situations, suffocation [35]. Although modern air conditioning systems are designed to minimize these risks and regular maintenance significantly reduces the likelihood of leaks, the closed and isolated nature of the station requires that any incident be treated extremely seriously.The probability of this risk is classified as unlikely; however, due to the potential harmful effects, which include death, serious injury, and irreversible damage, it is categorized as a medium-level risk. This highlights the need for continuous monitoring and rigorous preventive maintenance to ensure crew safety. It is essential to establish a protocol for the maintenance and cleaning of air conditioning units.

It is important to note that while some chemical hazards are part of analog training – preparing crew members to deal with possible exposures on real missions to Mars – others, such as refrigerant leaks, should be eliminated whenever possible as they do not add value to the experience. training and may compromise the safety of participants. The moderate risk rating reflects this duality, highlighting the importance of a robust preventative approach to ensure the safety and effectiveness of missions at the analogue station.

##### 4.1.2.3 Mechanical or accident risk

The Mars Lab workshop has a variety of tools distributed in various parts of the compartment. Improper handling of these tools can result in bruises, fractures and other serious injuries. Although the probability of accidents occurring is low, due to crew members limited contact with tools (classified as level 1), the severity of possible injuries is high (classified as level 3). Therefore, this risk is considered medium level. To mitigate these risks, it is essential to implement a protocol that centralizes the storage of tools in a single designated location, with access restricted to authorized people. Furthermore, the use of appropriate PPE, such as gloves, safety glasses and appropriate footwear, is essential when handling these tools. Regular maintenance and inspection of work areas are necessary to identify and eliminate mechanical hazards.

Furthermore, the possible presence of venomous animals within the station base presents an additional risk of accidents. Bites from scorpions, spiders and snakes, for example, can cause local reactions such as pain, edema and numbness, hemorrhagic reactions and vegas (vomiting and diarrhea), in addition to kidney and respiratory failure [36]. The state of Rio Grande do Norte has a high rate of accidents caused by venomous animals. Searches for Barbosa (2015) reported that, from 2007 to 2011, there were 15,694 accidents. As for the causative animal, 65.4% of recorded accidents were caused by scorpions, 13.5% caused by snakes, 5.2% caused by bees, 3.8% caused by spiders, 1% caused by caterpillars and 6.4% caused by other animals such as fish, ants, wasps and beetles. In 4.7% of accidents, the animal caused was not identified [36]. In cases of more serious accidents, there is a health post located 16 minutes from the station, as shown in Figure 4, which can provide additional medical support if necessary. However, if the use of anti-venom serum is necessary, the space analog station strategy involves activating local health units and SAMU. Considering that SAMU can take time to arrive in rural areas, the response time increases significantly, especially for transport to the nearest capital, Natal, which is approximately 95 km from the station. This delay constitutes a complication that further increases the risk to the lives of the crew. Due to the possible presence of venomous animals at geographic area where the station operates, especially scorpions, and the high severity of the risk, which can result in deaths, serious injuries and irreversible damage, this risk was classified as high.

**Fig. 4.**
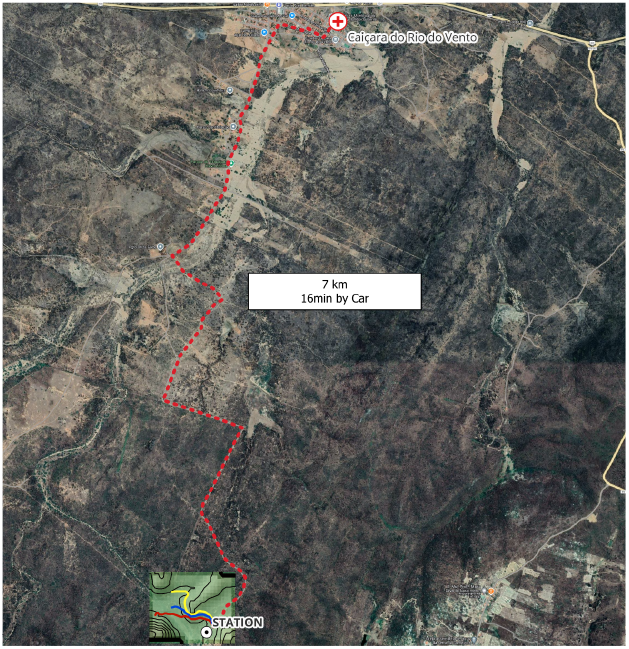
Distance from Habitat Marte to the nearest health center. Source: Author’s own elaboration.

To minimize this risk, it is essential to develop regular inspections in vulnerable areas, such as warehouses, dormitories and tool storage areas. Regular pest control during the season is also an important preventive measure to control the presence of these animals. Furthermore, it is necessary to train the crew to identify and deal with these animals safely, in addition to providing specific antidotes and first aid kits for bites.

It is important to distinguish between risks that must be eliminated completely and those that, although minimized, are part of analogous training. Risks such as lack of organization and inadequate centralization of tools, incorrect storage of heavy objects, and the entry of venomous animals into the base must be eliminated, as they do not contribute to training and can compromise crew safety. On the other hand, the safe handling of heavy tools with PPE and familiarity with working in environments where there is a risk of falling objects are part of the essential training for missions to Mars, preparing crew members to operate safely in adverse conditions.

##### 4.1.2.4 Physical Risk

The station, located in a region with an arid and dry climate, naturally faces extremely high temperatures, which poses a significant physical risk, such as dehydration, heat exhaustion, or heatstroke. The annual average air temperature is 27.6 ºC, with a peak of 29.2 ºC in October during the dry season, and a minimum of 25.9 ºC in June-July 2003, shortly after the end of the rainy season [37].

However, the station is equipped with air conditioning systems that keep the internal environment safe and comfortable, significantly reducing this risk. The presence of thermal blankets in the station’s insulation also helps maintain adequate temperatures and protects crew members from extreme heat. Therefore, the risk of heat exposure is classified as low, given its unlikely probability of occurrence—relevant only in the event of air conditioning system failure—and its severity limited to mild health disturbances. Additionally, the station is equipped with devices such as refrigerators, air compressors, and generators, which can produce vibrations and noise [38]. These factors may cause discomfort, stress, and hearing fatigue. The noise and vibration risk is considered low, and since the station’s equipment does not generate significant noise, the use of ear protection is not always necessary. Proper maintenance and strategic placement of devices can further reduce exposure, especially during rest periods.

In the context of a real mission to Mars, the crew will be exposed to constant noise as well as exposure to microgravity during the space travel and hypogravity when already in place on the red planet. The latter can alter biomechanics which, in addition to sarcopenia and osteopenia, can generate changes in the spinal column, generating pain and disc inflammation [39]. Therefore, while physical risks that can seriously impact the health and integrity of crew members must be eliminated, such as prolonged exposure to unmitigated vibrations, others, which simulate real mission conditions, must be minimized, but maintained as part of training and data collection. These practices must be accompanied by approval from the Ethics Committee to ensure the safety and well-being of participants.

##### 4.1.2.5 Ergonomic Risk

Prolonged exposure to the environment, especially in static or uncomfortable positions for long periods, requires frequent breaks for movement, stretching and muscle relaxation exercises. Inside the station, crew members face ergonomic challenges due to the confined space and the design of the work and rest areas. Confined spaces, caused by small compartments that limit freedom of movement, can be optimized through interior design that maximizes available space, use of multifunctional furniture and efficient arrangement of space to allow for easy movement. The risk associated with confined spaces is moderate, as although they can cause discomfort and limit mobility, design solutions and the use of suitable furniture can effectively minimize these problems. Prolonged postures, resulting from long periods of sitting or standing during work or rest, can be mitigated with adjustable chairs and tables, recommending regular breaks for stretching and changing positions, and promoting a routine that includes periods of physical activity and stretching. Prolonged postures present a moderate risk, as, without appropriate breaks and ergonomic adjustments, they can lead to significant discomfort and musculoskeletal problems, however, they are relatively easy to mitigate with adequate ergonomic practices [40; 41].

Manipulation of equipment and tools, caused by the frequent use of hand and electronic tools, can be facilitated with ergonomically designed tools, adequate training in the use of equipment and the provision of supports or accessories that reduce physical effort [42]. The risk associated with handling equipment and tools is moderate to high, as if they are not used correctly or are poorly designed, they can lead to repetitive strain injuries or more serious accidents, requiring careful attention to handling practices and the design of tools. Visual stress, the result of prolonged use of computer screens and other visual devices, can be prevented with adequate lighting, monitors with adjustable brightness and contrast, and regular eye rest breaks. Visual stress is classified as a low to moderate risk because, although it can cause discomfort and eye fatigue, its consequences are generally less serious and can be easily managed with good lighting practices and regular breaks. Visual stress can also negatively affect motor skills when performing tasks [43], with galvanic vestibular electrostimulation emerging as a countermeasure for this [44].

Among the ergonomic risks, psychosocial risks stand out, which can manifest themselves significantly in stations similar to Mars. In these isolated and confined environments, crew members are exposed to conditions of sensory deprivation, prolonged isolation from their social and family circles, and the constant pressure of performing critical tasks. Studies show that these factors can lead to the development of chronic stress, anxiety, depression and mental fatigue, compromising both the well-being of individuals and the overall performance of missions [45]. During the stay at the station, some strategies were implemented to mitigate these risks, such as the practice of meditation and Mindfulness, which demonstrated effectiveness in reducing stress and improving mental focus [46]. These techniques promote relaxation, emotional control and help strengthen crew members’ resilience in the face of the challenges of the extreme environment. According to the station manager, this practice takes place before the crew goes to sleep, providing relaxation after a day of activities. Although psychosocial risks are especially significant in contexts of confinement and isolation, the station organization adopted proactive measures to minimize their impacts. Psychological stress in isolation environments, such as the Habitat Marte space analog station, can be intensified by confinement and environmental pressure, often resulting in interpersonal conflicts and communication problems. Increasing the number of participants could help reduce the feeling of isolation, creating a more dynamic and collaborative environment. Given that this risk is classified as medium probability and high severity, it was considered a level 2 risk, requiring continuous monitoring and effective minimization strategies to preserve mental health and mission success.

### 4.2 Underwater Extravehicular Activities

#### 4.2.1 Description of activities carried out

The pool is made with fiber and measures 5m x 2.80m. It is located in the external area of the station base and is used for reduced gravity simulation training, essential for the preparation of crew members. Activities include handling tools, physical exercises, buoyancy and microgravity training, as well as underwater psychological relaxation exercises, where crew members are submerged with breathing apparatus and ambient sounds in headphones.

#### 4.2.2 Identification of critical points

During Mission 134 (December/2022), the presence of wasps was identified (*Hymenoptera: vespidae*) and bees (*Anthophila*) during the execution of UEVA (Figure 5), representing a risk of accidents. Although no accidents have occurred, the risk remains, especially considering that these insects tend to surround the pool, with the possibility of being accidentally sucked in by respirators used during underwater activities. Additionally, bee stings can cause serious allergic reactions, such as difficulty breathing, itching, redness, increased heart rate, and dizziness [47], representing a medium to high risk, depending on the individual sensitivity of the crew, the density of insect colonies and the effectiveness of preventive measures.

**Fig. 5.**
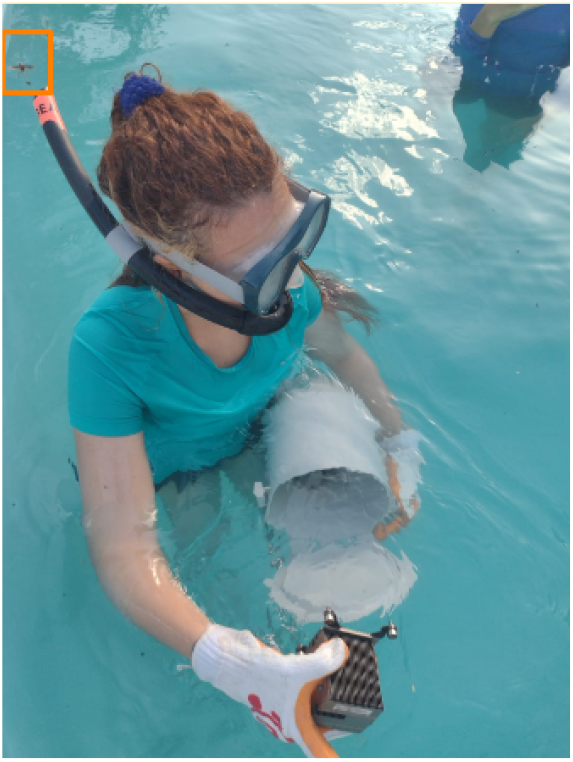
Application of UEVA, where the author is handling instruments underwater, simulating what the movement of these objects would be like in microgravity. Note in the upper left corner, highlighted in orange, the presence of a wasp near the analogue astronaut’s respirator. Source: Author’s own elaboration.

Additionally, an emotional imbalance was noted in participants who had fears and/or phobias related to these insects, compromising their immersion in the experience and their well-being during the activity, which was classified as an ergonomic/psychosocial risk. This risk can be categorized from low to medium, depending on the individual level of fear of each participant, and can lead to mild symptoms of discomfort up to paralysis due to a panic attack [48].

To mitigate these risks, it is essential that crew members receive adequate training in first aid and accident prevention before the mission begins, in addition to strictly adhering to safety protocols. The presence of nearby emergency response bases is highly recommended, ensuring rapid response in the event of incidents. As there will be no exposure to insects on manned missions to Mars, this concern should be eliminated in this UEVA experience. Therefore, it is recommended to use protective nets around the pool to minimize this risk. Furthermore, as demonstrated by LIMA (2019), there are several ecological methods to ward off insects, using plants native to Brazil, such as the açaí tree (*Euterpe oleracea* Mart.) [49]. According to the study, burning bunches of this plant produces effective smoke to repel insects. Therefore, we suggest planting açaí palms and using this practice during UEVA. Considering that the pool uses chemical substances for treatment, it is essential to develop strict monitoring of the water to avoid contamination and adverse reactions. The use of these products must follow ANVISA guidelines, ensuring safe control of permitted concentrations [50].

### 4.3 Risk Assessment Map: IVA and UEVA

This item presents the risk map developed for the analogous station base, highlighting the areas of greatest vulnerability and the risks previously discussed. The base of the station has structures for activities intraveiculares and underwater extravehicular vehicles. Due to the proximity of these activities, both were represented on the same map (Figure 6 and Figure 7).

**Fig. 6.**
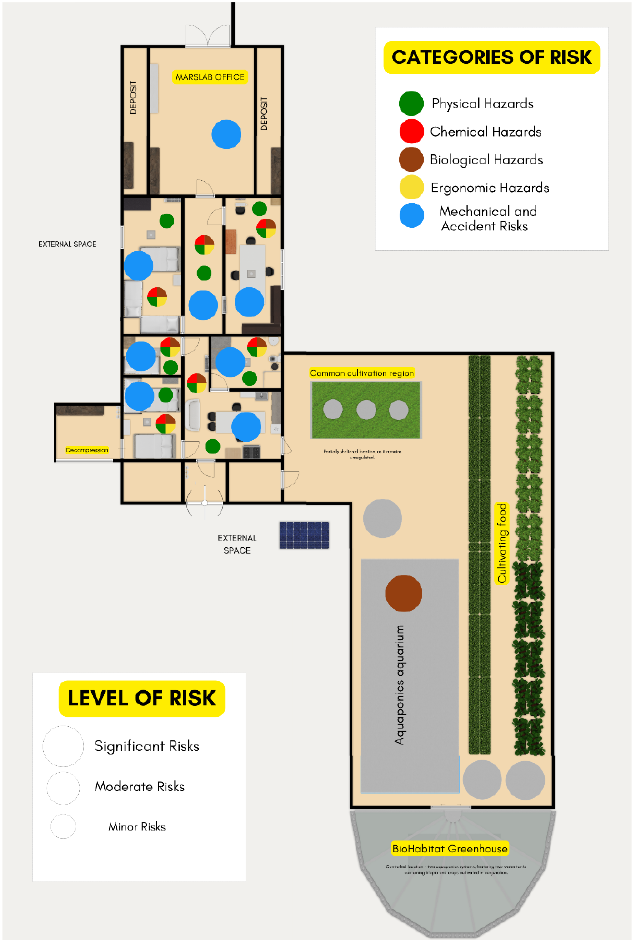
Risk map of the base of the Habitat Marte analogue station. Source: Author’s own elaboration.

**Fig. 7.**
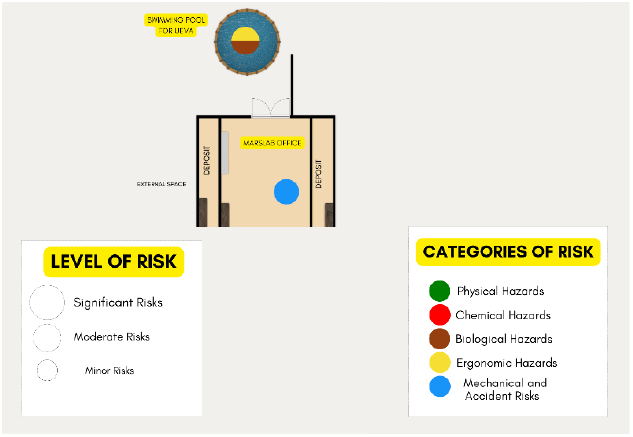
UEVA risk map. Source: Author’s own elaboration.

### 4.2 Extravehicular Activities

EVAs include visits to Pico do Cabugi, DeltaCave, LavaCave and Lake Ceres. The PPE provided for these activities are helmets, clothing and gloves.

#### 4.2.1 Description of activities carried out

#### 4.2.1.1 Cabugi Peak

In this activity, crew members follow a previously mapped route that includes walking challenges and overcoming terrain with loose rocks and gravel to reach Pico do Cabugi, an inactive volcano in Brazil. The journey starts at the base station, located at a distance of 43.6 km from Pico, where the crew receives instructions for this EVA and recommendations on what to take, such as water and sunscreen, in addition to stretching sessions.

After this moment, the crew is moved by car to the base of the peak and then begins the journey on foot. The route includes a gradual walk around the base of the volcano, where crew members carry out soil assessments and collect geological samples. Pico do Cabugi, almost 600 meters high, presents a significant variation in relief (Figure 11), providing a challenging and ideal terrain for simulating space missions in rugged environments. However, the team reaches approximately 350 meters of altitude, without completing the climb to the peak.

#### 4.2.1.2 DeltaCave

The extravehicular activity in DeltaCave aims to reach a cave formed by two rocks that take on a triangular shape. This destination is difficult to access, located at the top of a geological formation with a steep slope and high variation in declivity along the route. Due to the slope of the terrain, the route is carried out with the support of ropes to guarantee the safety of the crew. It is important to highlight that, at the time of writing this article, this EVA was temporarily inaccessible due to the risks associated with accessing the location.

#### 4.2.1.3 LavaCave

The Lava Cave Habitat aims to simulate a lava tube, considered a possible location for human life and settlement on Mars. Located 600 meters from the analogous space station Habitat Marte, it is a strategic location for simulating space missions and training astronauts in extreme environments. The activities carried out at Lava Cave include improvements of the facility, food preparation, assessment of the use of facilities and sleep quality.

#### 4.2.1.4 Ceres Lake

Observation of the night sky takes place in a structure located next to Ceres Lake. To reach the observation point, the crew takes a walk just before sunset. Preparation for this EVA includes selecting essential equipment, such as flashlights and gloves, to ensure a safe return to the station after the sun sets, when natural light is non-existent.

#### 4.2.2 Identification of critical points

#### 4.2.2.1 Biological Risk

Throughout all EVAs, crew members encounter significant biological risks, primarily associated with the presence of disease vectors. During manned mission 134, vectors such as bats and mosquitoes were observed on the routes to the DeltaCave and LavaCave caves. These vectors have the ability to transmit pathogens responsible for skin infections and other diseases, posing a threat to the health of crew members [51; 52]. As they are likely to occur occasionally and can cause deaths, serious injuries and irreversible damage, this risk was classified as high. For effective mitigation, it is essential to use appropriate protective clothing, apply repellents and carry out thorough inspections of activity areas before starting EVAs. The transmission of diseases through these vectors is considered indirect, as it involves intermediaries such as the vectors themselves or toxic organisms present in the environment.

In addition to the preventive measures mentioned, vaccination against rabies can be an important strategy to protect crew members, especially due to the presence of bats, which are known transmitters of the rabies virus. Implementing infection control and containment practices, such as the use of appropriate Personal Protective Equipment (PPE) and disinfection procedures, is essential to minimize these risks. Biological risks identified during EVAs are classified as level 3, indicating a moderate individual risk. Although these risks do not pose a significant threat to the community, they require the application of prophylactic and therapeutic measures, as well as the adoption of strict infection control practices to ensure the safety of crew members. These risks presented must be eliminated as they do not add to the qualification and training of the similar astronaut, as they are not risks that would be present in a real mission to Mars.

#### 4.2.2.3 Mechanical or accident risks

Crew members face numerous possibilities of mechanical and accident risks during EVAs, many of which are exacerbated by the natural conditions surrounding the station. Among the main risks are those associated with venomous animals, which are possibly seen, especially in the Lago Ceres area, due to the nocturnal habits of these animals and the lack of light during the return to base. Studies conducted by TAVARES et al., 2020 in the state of Rio Grande do Norte indicate that the majority of accidents involving venomous animals are caused by scorpions. However, snakebites, although less frequent, present a higher risk of fatality. Furthermore, there have been recorded incidents involving spiders, caterpillars and bees, which highlights the diversity of dangers to humans present in the environment [53]. Contact with these animals can result in bites which, if not treated promptly, can be fatal.

The risk is amplified by the difficulty in accessing emergency services and the absence of antivenom at the station. Given the average probability of accidents involving venomous animals and the high severity of the consequences, which may include the possibility of death, this risk was classified as high. After consultation with the manager of the Habitat Marte Analog Station, it was found that there are safety procedures in place. In the event of bites, the situation becomes critical due to the lack of antivenom, requiring the immediate transportation of the crew member to Natal, the state capital. It is recommended that a rapid response protocol be implemented that includes identifying the animal if possible, cleaning the affected area with soap and water, applying ice to control swelling, and continuing monitoring for signs of allergic or allergic reactions and infections. If there are severe symptoms, the evacuation protocol must be immediately followed for specialized medical care [54]. Protocols at Habitat Marte are currently being updated during this research.

In this specific mission, accidents involving cactus and other thorny plants present in the path of extravehicular activities were also observed, which can cause scratches, bleeding and even more serious injuries [54; 55]. Thorny plants, such as cactus and nettles, also pose mechanical risks. Inadvertent contact with these plants, especially nettles, can cause scratches, skin irritation and more serious injuries [57], especially in rough terrain and during exploration activities. To mitigate these risks, it is essential to flag areas with a high concentration of these plants and provide adequate protective clothing to crew members. Although most symptoms are treatable and the frequency of these accidents is low, the availability of ointments to relieve irritation is recommended. Based on the rarity of accidents and the possibility of certain serious injuries, this risk was considered medium level.

The risk of accidents during EVAs is also influenced by geological and climatic conditions. The route to Pico do Cabugi presents uneven terrain, with slopes varying between 45% and 75% (Figure 11), slippery surfaces and loose rocks, which increases the risk of falls and slipping (Figure 8). In the DeltaCave and LavaCave regions, although the risk is relatively lower, there is still danger due to the variation in relief and the presence of unstable geological formations. To mitigate these risks, it is crucial that crew members use appropriate Personal Protective Equipment (PPE), such as helmets, hiking boots, trekking poles and ropes to assist with ascent in steep areas. Considering that the probability of accidents, such as slips and trips, is occasional and that such incidents can result in serious injuries, irreversible damage or even death, this risk was classified as high. Furthermore, the collapse of structures in narrow passages, especially in LavaCave, represents a significant concern given the risk of collapses due to geological instability. Although the station provides general instructions to crew members, it is recommended that more in-depth practical training be implemented, including climbing techniques, use of knots, safety equipment and appropriate practices for climbing in natural or structural environments.

**Fig. 8.**
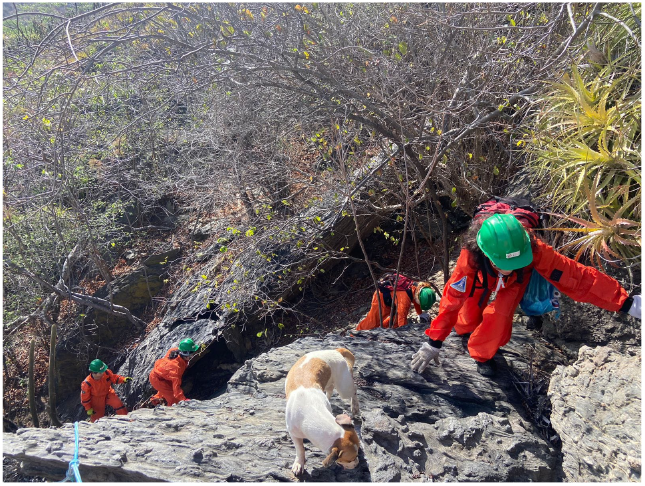
Path taken for DeltaCave extravehicular activity. Source: Author’s own elaboration.

Brazil is a country frequently ravaged by large-scale forest fires that cause immense ecological and financial damage [58]. In Rio Grande do Norte, the occurrence of fires, whether controlled or not, is common. Due to the climatic seasonality characteristic of Brazil’s semi-arid region, in the dry season this region presents high temperatures, low relative humidity and increased wind speed. [58]. Therefore, creating evacuation plans is essential during EVAs. In addition, it is recommended to carry out periodic training to prepare in case of fires, as well as monitor weather conditions and the presence of hot spots in the vicinity of the station. The adoption of risk mitigation practices, such as the creation of safety areas and the use of firefighting tools, is essential to reduce the vulnerability of crew members to this type of threat. In cases of burns, it is essential that the station has an emergency protocol, which contains instructions for identifying the severity of the burn and appropriate first aid measures. Due to the low probability but high severity of this risk, which can result in serious burns and even fatalities, it has been classified as medium level.

Figure 9 illustrates the fires during Mission 134. Although these fires are not located within the Habitat Marte area, several hot areas in the region highlight the potential for recurring fire incidents in the state. This evidence reinforces the urgent need to create and implement specific security protocols to mitigate this risk. Recommended protocols include the development of a detailed evacuation plan with escape routes and safe meeting points, as well as regular training to simulate fire situations and evacuation practices. Additionally, it is crucial to continuously monitor weather conditions and hot spots near the station to enable a rapid response. The creation of safety areas around the station and the provision of adequate firefighting tools are also essential to protect the integrity of the station and the safety of the crew.

**Fig. 9.**
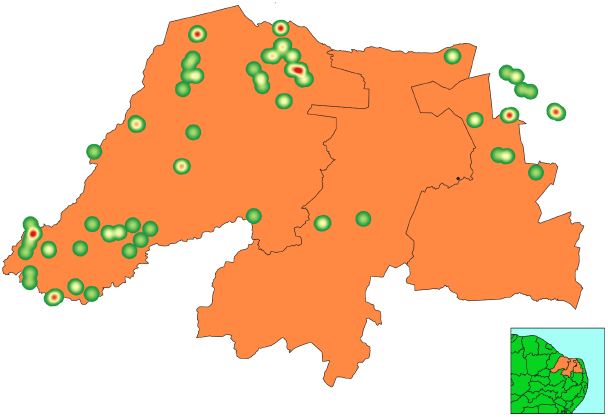
Kernel map (heat) for visual identification of “hot areas”, that is, with a high concentration of fires, during the period of December 2022. Data source: BDQueimadas of INPE’s Fire Monitoring Program. Source: Author’s own elaboration.

Certain risks, such as accidents related to contact with plants and animals and forest fires, must be eliminated, as they have no similarity with the context and reality of Mars. On Mars, natural fires would not occur due to the absence of sufficient oxygen to sustain combustion, low atmospheric pressure, lack of vegetation or flammable materials, and the planet’s cold and dry climate [59; 60]. Therefore, these risks do not represent a threat and could compromise the safety of those involved in the immersion if maintained without purpose. However, risks associated with geological conditions must be maintained, but mitigated, so that the physical integrity of members is preserved. Implementing standardized practices and security protocols is crucial to ensuring these risks are addressed effectively.

#### 4.2.2.4 Physical Risks

During this mission, several physical risks associated with EVAs, including exposure to intense heat, dust and high solar radiation. The dust found is characteristic of hot and arid environments and can be carried by the wind and contain fine particles that, when inhaled, can cause respiratory problems and eye irritation. These conditions make Habitat Mars an ideal place for studies and simulations of Martian environments, enabling the observation of processes similar to those on the red planet. Although there is a high probability of exposure to dust, health impacts can be mitigated with the appropriate use of Personal Protective Equipment (PPE). Therefore, the risk was classified as medium, indicating that, although moderate, it can be controlled with effective preventive measures. However, it is essential to maintain constant monitoring to avoid long-term health complications.

One of the risks most highlighted by the crew during this mission was exposure to heat. Prolonged exposure to heat in semi-arid climates poses a significant health risk, with the potential to cause heatstroke, exhaustion, and, in severe cases, complications such as respiratory distress syndrome [61]. Mitigation involves wearing light, protective clothing, staying hydrated, taking frequent breaks, and planning activities during cooler times, such as early in the morning or late in the afternoon. Intense solar radiation, also classified as high risk, can cause burns, damage to the skin and increase the risk of cancer [62]. To reduce this risk, it is recommended to use sunscreen, hats, clothing with UV protection and limit sun exposure. The use of wearable devices that monitor vital signs, such as body temperature, heart rate and blood oxygenation, can warn of dehydration or thermal exhaustion, allowing immediate interventions. Staff training on the signs of heatstroke and the proper use of PPE are also essential to avoid complications.

Another important risk is dehydration, amplified by the combination of high temperatures and low humidity, which can result in rapid loss of body fluids and thermal shock. Furthermore, the variation in altitude and slope on Pico do Cabugi, where the crew covers between 300 and 400 meters of the total 565 meters of the Pico (Figure 11), can cause physiological changes, such as extreme tiredness and consequent shortness of breath [63]. Although altitude itself is not enough to cause hypoxia, the effects of the low humidity and temperature variation of the semi-arid climate can result in symptoms such as dizziness, fatigue and disorientation, exacerbating the risk of heatstroke and dehydration. Low humidity conditions also posed a significant risk during the mission, contributing to dry skin and mucous membranes, which increases the chance of irritation and infection. Prevention involves constant hydration, the use of moisturizing creams and monitoring weather conditions, as low humidity can go unnoticed or be confused with other symptoms [64]. Despite the high probability of exposure to heat, low humidity and altitude variations, especially at Pico do Cabugi, the risk was classified as medium level, due to the possibility of effective control through preventive measures and continuous monitoring.

To mitigate these risks, it is essential that the station provides protocols and prior physical training, aiming to increase resistance to the semi-arid climate, characterized by high temperatures and environmental pressures. Lack of prior conditioning, both musculoskeletal and aerobic, is associated with a greater risk of injuries, such as sprains, strains and muscle tears [65]. Furthermore, the use of appropriate equipment and regular monitoring of crew members’ physical conditions, before and during EVAs, are essential to ensure safety. Partnerships with centers specializing in climbing training are also recommended, providing more robust and specific preparation for the physical challenges faced by crew members.

These environmental conditions and physical challenges reinforce the importance of the Habitat Marte space analog station as an ideal analogue for manned missions to Mars, enabling the observation and analysis of processes that may occur on Mars and the implementation of rigorous and well-planned safety practices both on Earth and in space. In this aspect, resistance exercises (applied with elastic bands or springs) are essential for maintaining the locomotor system in good working order, in addition to being an efficient countermeasure both in analogous space and in microgravity [66].

For effective training for missions to Mars, it is crucial to focus on risks that reflect the conditions and challenges astronauts are likely to encounter on the red planet. Risks such as exposure to heat, dust, solar radiation, dehydration and altitude variations must be mitigated with appropriate measures, but maintained, to ensure that crew members are well prepared.

#### 4.2.2.5 Ergonomic Risks

Ergonomic risks in EVAs are significant and range from moderate to high, due to the potential to cause physical discomfort and injuries to crew members. During EVAs, crew members are often exposed to long walks and repetitive movements, which can lead to repetitive strain injuries and muscle fatigue. Constant use of the same muscle groups during these activities increases the risk of developing musculoskeletal problems, such as tendinitis and strains [68]. To mitigate these risks, it is essential to implement an effective ergonomic prevention program. The first preventive measure involves performing warm-up and stretching exercises before and after EVAs, preparing muscles and joints for physical activity and reducing the likelihood of injuries. Furthermore, it is essential to schedule regular rest breaks during activities, allowing crew members to recover and reduce accumulated muscle tension. Rotating tasks between crew members is also an important strategy to avoid repetitive use of the same muscle groups. Alternating activities distribute the physical load more evenly, reducing the risk of repetitive strain injuries.

When transporting heavy or bulky equipment during EVAs, it is recommended to use ergonomic backpacks that distribute the weight evenly, minimizing stress on the back and shoulders. Additionally, balanced load distribution among crew members and the use of transport devices such as carts can reduce the need to manually carry excessive weight. It is also suggested to implement protocols with practical training on the notions of hiking with heavy luggage, describing how to adjust backpacks and appropriate weight distribution. Another relevant ergonomic risk is incorrect postures adopted during tasks, often due to the configuration of the terrain or the nature of the activities. Preventing these forced postures can be achieved through prior stretching that increases crew members’ flexibility and prepares them for more demanding movements. The use of adjustable equipment, which allows the maintenance of an adequate posture, is essential. For example, devices that adjust working height can minimize the need to bend, squat or twist the body. Careful planning of activities is also crucial, ensuring that tasks are organized in a way that minimizes forced postures as much as possible.

During the journey to EVAs, in Mission 134, several psychosocial challenges were identified. Participants faced significant emotional and psychological impacts, mainly due to the constant state of alert caused by the possibility of accidents and the risk of unexpected situations, such as the appearance of venomous animals or abrupt changes in the environment. This constant level of vigilance can deteriorate well-being, generating anxiety and stress, which, in turn, increases the likelihood of errors and compromises the safety of operations.

Faced with these challenges, it is recommended that, in addition to practicing Mindfulness before bed, it is also incorporated before EVAs. Mindfulness, by reducing excessive alertness and anxiety, can contribute to reducing heart rate and promoting a calmer, more focused mind [69]. This can not only reduce the occurrence of accidents, but also improve performance in simulated missions. Additionally, the application of controlled breathing techniques would complement these benefits [69], helping to regulate stress and mentally and physically prepare participants for EVAs. In the context of a space analog mission, certain ergonomic risks, such as the transport of heavy equipment and the need for varied postures, are part of the training and must be minimized, but maintained to simulate the real conditions of a space mission.

However, risks that can be eliminated, such as a lack of regular breaks or inappropriate use of equipment, must be addressed with appropriate preventative measures to protect the health and well-being of crew members. Although ergonomic risk can have serious impacts, its likelihood and severity can be significantly reduced through rigorous application of prevention protocols and adequate training. Therefore, the risk is classified as level 2, indicating the need for continuous mitigation measures to ensure the safety and well-being of crew members.

#### 4.2.3 Risk Assessment Map: EVA

In this section, risk maps associated with extravehicular activities will be presented. Figure 10 illustrates the different risks mapped along the routes of activities carried out in the regions DeltaCave, LavaCave and Lago Ceres. Figure 11, in turn, highlights the risks identified during the operations carried out at Pico do Cabugi.

**Fig. 10.**
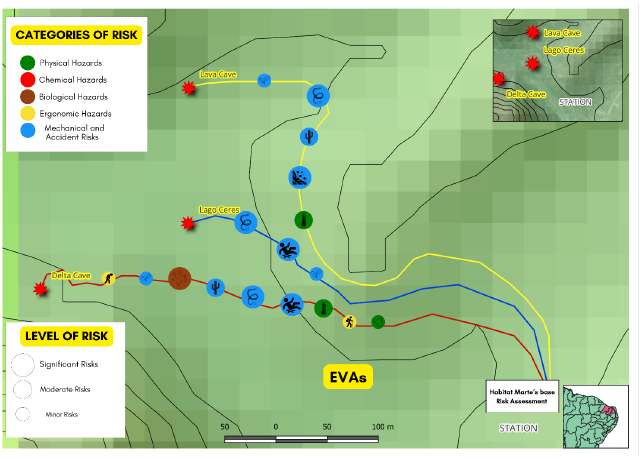
Risk Assessment Map highlighting the main hazards associated with extravehicular activities LavaCave, DeltaCave and Lake Ceres of the Mars analogue station, Habitat Marte. Source: Author’s own elaboration.

**Fig. 11.**
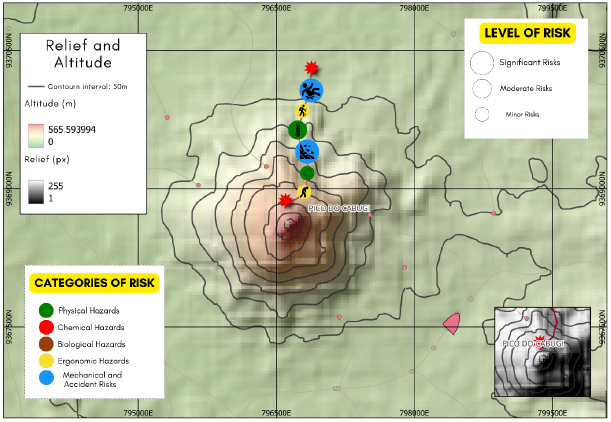
Risk map highlighting the main dangers associated with extravehicular activities carried out at Pico do Cabugi. Source: Author’s own elaboration.

## 5. Conclusions

The comprehensive mapping of occupational risks at the Habitat Marte analog station is essential for ensuring the safety and wellbeing of the crew during missions. This study identified a range of risks, including biological, chemical, mechanical, physical, and ergonomic hazards, which are crucial for anticipating and mitigating potential incidents in extreme environments. Defining these risks not only protects the participants but also enhances the overall mission success by preparing crew members for the conditions they might face during real space missions.

Moreover, it is crucial to establish clear objectives for analog missions. These missions should primarily serve as a platform for training and certifying analog astronauts, preparing them for the challenges of long-duration space travel and life on extraterrestrial bodies like Mars. Some risks, such as those involving biological and mechanical hazards, must be carefully managed to avoid compromising safety without adding value to the training. However, controlled exposure to risks that mirror real space missions should be maintained as part of the selection process and training regimen. Based on the observations in the case study and the interview applied with the Habitat Marte manager, a questionnaire was proposed to be applied in future missions, which addresses issues related to satisfaction with general safety at the station, including the effectiveness of training, the adequacy of PPE, the clarity of emergency protocols, the quality of remote medical monitoring and suggestions for improving safety in future missions. This questionnaire aims to achieve continuous improvement in station safety and the adequacy of safety protocols, ensuring that preventive measures are constantly improved and that crew members’ needs and concerns are duly addressed.

Although there are already more than 200 protocols in operation at the Habitat Marte station, recently identified risks show that it is still necessary to develop new procedures and improve existing ones. While these current protocols are effective in mitigating many dangers, they do not fully cover all potential situations. This need reinforces the importance of continuous reviews and the development of additional protocols that can address emerging scenarios, ensuring even more comprehensive safety for participants.

## Data Availability

All data produced in the present work are contained in the manuscript

https://iafastro.directory/iac/proceedings/IAC-24/

## Acronyms/Abbreviations

(CIPA): Internal Accident Prevention Commission
(NR): Regulatory Standard
(DEM): Digital Elevation Model
(NASA): National Aeronautics and Space Administration
(EVA): Extravehicular Activity
(UEVA): Underwater Extravehicular Activity
(IVA): Intravehicular Activity
(PPE): Personal Protective Equipment
(READ): Repetitive strain injuries
(WMSD): Musculoskeletal diseases related to work
(m): Meters
(km): Kilometers
(PRA): Probabilistic Risk Assessment
(GIS): Geographic Information Systems
(SRTM): Shuttle Radar Topography Mission
(INPE): National Institute for Space Research
(IBGE): Brazilian Institute of Geography and Statistics
(SAMU): Mobile Emergency Care Service
(ANVISA): National Health Surveillance Agency

## References

[1] PROENÇA, Ana Cláudia Taveira. Mapa de Riscos da FEUP-Revisão e aplicação a casos de estudo. 2017.

[2] Nascimento, Marta Oliveira; ARAÚJO, Giovana Fernandes. Riscos ocupacionais dos profissionais de enfermagem atuantes no SAMU 192. ID on line. Revista de psicologia, v. 10, n. 33, p. 212–223, 2017.

[3] Linck, Evan et al. Evaluation of a Human Mission to Mars by 2033. Washington, DC: IDA Science and Technology Policy Institute, 2019.

[4] Paris, Antonio. Physiological and psychological aspects of sending humans to Mars: Challenges and recommendations. Journal of the Washington Academy of Sciences, v. 100, n. 4, p. 3–20, 2014.

[5] Manzey, Dietrich. Human missions to Mars: new psychological challenges and research issues. Acta Astronautica, v. 55, n. 3-9, p. 781–790, 2004.

[6] Stoker, Carol R. et al. Mineralogical, chemical, organic and microbial properties of subsurface soil cores from Mars Desert Research Station (Utah, USA): phyllosilicate and sulfate analogues to Mars mission landing sites. International Journal of Astrobiology, v. 10, n. 3, p. 269–289, 2011.

[7] Binsted, Kim et al. Human factors research as part of a Mars exploration analogue mission on Devon Island. Planetary and Space Science, v. 58, n. 7-8, p. 994–1006, 2010.

[8] Souto, Sofia; ALABART, Joan; Costa, Hugo André. An overview of space analogues in Portugal. 2023.

[9] De Souza, Davi Alves Feitosa; DE REZENDE, Julio Francisco Dantas; Bentes, Anibal Picanço. A exploração espacial no cenário educacional, tecnológico e sustentabilidade no Brasil: o caso da estação espacial análoga habitat marte Space exploration in the educational, technological and sustainability scenario in Brazil: the case of the mars habitat analog space station. Brazilian Journal of Development, v. 8, n. 2, p. 14919–14934, 2022.

[10] Rai, B., & Kaur, J. (2012). Human Factor Studies on a Mars Analogue During Crew 100b International Lunar Exploration Working Group EuroMoonMars Crew: Proposed New Approaches for Future Human Space and Interplanetary Missions. North American Journal of Medical Sciences, 4, 548–557. 10.4103/1947-2714.103313.

[11] Sidorov, A., Bogdanov, A., Medvedeva, Y., & Filippov, A. (2021). Occupational Risk Determination Using the Integral Assessment of Working Conditions Methodology., 88–93. 10.24000/0409-2961-2021-3-88-93

[12] Rodrigues, Luciano Brito; SANTANA, Nívio Batista. Identificação de riscos ocupacionais em uma indústria de sorvetes. Journal of Health Sciences, v. 12, n. 3, 2010.

[13] Kim, J., & Lee, B. (2007). Risk Factors, Health Risks, and Risk Management for Aircraft Personnel and Frequent Flyers. Journal of Toxicology and Environmental Health, Part B, 10, 223–234. 10.1080/10937400600882103.

[14] Patel, Z., Brunstetter, T., Tarver, W., Whitmire, A., Zwart, S., Smith, S., & Huff, J. (2020). Red risks for a journey to the red planet: The highest priority human health risks for a mission to Mars. NPJ Microgravity, 6. 10.1038/s41526-020-00124-6.

[15] Baran, R.; Marchal, S.; Garcia Campos, S.; Rehnberg, E.; Tabury, K.; Baselet, B.; Wehland, M.; Grimm, D.; Baatout, S. The Cardiovascular System in Space: Focus on In Vivo and In Vitro Studies. Biomedicines 2022, 10, 59. 10.3390/biomedicines10010059

[16] Bonani R; Cariati I; Marini M; *et. al*. Microgravity and Musculoskeletal Health: What Strategies Should Be Used for a Great Challenge?, Life 2023. 13, 1423

[17] Bizzarri, Mariano et al. Journey to Mars: A Biomedical Challenge. Perspective on future human space flight. Organisms. Journal of Biological Sciences, v. 1, n. 2, p. 15–26, 2017.

[18] BRASIL. Portaria n. 3.214, de 08 de junho de 1978. Ministério do trabalho e emprego. Norma Regulamentadora 09 - Avaliação e Controle das Exposições Ocupacionais a Agentes Físicos, Químicos e Biológicos, Brasília, DF, 2023.

[19] BRITO, Cássio Felipe Bandeira de. Investigação e Desenvolvimento de Metodologia de Avaliação dos Riscos Ergonômicos Psicossociais. 2022.

[20] Gabriel, G. et al. Future perspectives on space psychology: recommendations on psychosocial and neurobehavioural aspects of human spaceflight. Acta Astronautica, v. 81, n. 2, p. 587–599, 2012.

[21] Strangman, Gary E.; Sipes, Walter; Beven, Gary. Human cognitive performance in spaceflight and analogue environments. Aviation, space, and environmental medicine, v. 85, n. 10, p. 1033–1048, 2014.

[22] Mintus, Agata; ORZECHOWSKI, Leszek; Ćwilichowska, Natalia. Lunares analog research station—overview of updated design and research potential. Acta Astronautica, v. 193, p. 785–794, 2022.

[23] Xin, P., Khan, F., & Ahmed, S. (janeiro de 2017). Dynamic hazard identification and scenario mapping using Bayesian network. Process Safety and Environmental Protection, 105, 143155. doi:10.1016/j.psep.2016.11.003

[24] Cox, J. L. (2012). Evaluating and Improving Risk Formulas for Allocating Limited Budgets to Expensive Risk-Reduction Opportunities. Risk Analysis, 32(7), 1244–1252. doi:10.1111/j.15396924.2011.01735.x

[25] Mattos, Ubirajara A. de O.; FREITAS, Nilton Benedito B. Mapa de risco no Brasil: as limitações da aplicabilidade de um modelo operário. Cadernos de Saúde Pública, v. 10, p. 251–258, 1994.

[26] Ponzetto G. Mapa de Riscos Ambientais, São Paulo: Editora LTR, 3^ª^ Edição, 2010.

[27] Novello, Rosanna; NUNES, Rogerio da Silva; Marques, Roberto Salatiel Rodrigues. Análise de processos e a implantação do mapa de risco ocupacional em serviços de saúde: um estudo no serviço de hemoterapia de uma instituição pública federal. In: Congresso Nacional de Excelência em Gestão. 2011.

[28] Stamatelatos, Michael et al. Probabilistic risk assessment procedures guide for NASA managers and practitioners. 2011.

[29] Santos, Kamila Souza et al. Variabilidade temporal da cobertura da mata seca (caatinga) no estado do Rio Grande do Norte. Revista Ibero-Americana de Ciências Ambientais, v. 13, n. 8, p. 145–156, 2022.

[30] Silva, Maria Luana Oliveira et al. Áreas degradadas no Semiárido: Causas, situação e alternativas de recuperação. Ciências Rurais em Foco Volume 3, p. 22, 2021.

[31] DE Queiroz, Alexandre Cássio et al. Percepção dos Alunos da Área de Gestão Sobre a Relevância da Sustentabilidade Ambiental na Gestão Empresarial. Revista Controladoria e Gestão, v. 4, n. 2, p. 952–965, 2023.

[32] Wong, Wing C. et al. Preventing infectious diseases in spacecraft and space habitats. Modeling the Transmission and Prevention of Infectious Disease, p. 3–17, 2017.V

[33] Brasil. Ministério do Trabalho e Emprego. Norma Regulamentadora NR-24: Instalações Sanitárias e de Conforto nos Locais de Trabalho. Available at: https://www.gov.br/servidor/pt-br/siass/centrais_conteudo/manuais/nr-mte-24-instalacoes-sanitarias-e-de-conforto-nos-locais-de-trabalho.pdf. Accessed: September 18, 2024.

[34] Mori, J., & Smith, R. (2019). Transmission of waterborne fish and plant pathogens in aquaponics and their control with physical disinfection and filtration: A systematized review. Aquaculture. 10.1016/J.AQUACULTURE.2019.02.009.

[35] Mcfarland, M. (1992). Investigations of the environmental acceptability of fluorocarbon alternatives to chlorofluorocarbons.. Proceedings of the National Academy of Sciences of the United States of America, 89, 807–811. 10.1073/PNAS.89.3.807.

[36] Tsai, W. (2005). An overview of environmental hazards and exposure risk of hydrofluorocarbons (HFCs).. Chemosphere, 61 11, 1539–47. 10.1016/J.CHEMOSPHERE.2005.03.084.

[37] Barbosa, Isabelle Ribeiro. Aspectos clínicos e epidemiológicos dos acidentes provocados por animais peçonhentos no estado do Rio Grande do Norte. Revista Ciência Plural, v. 1, n. 3, p. 2–13, 2015.

[38] DA SILVA Santana José, Augusto; SILVA Souto Jacob,. Produção de serapilheira na Caatinga da região semi-árida do Rio Grande do Norte, Brasil. Idesia (Arica), v. 29, n. 2, p. 87–94, 2011.

[39] Huang, Y., & Griffin, M. (2014). The relative discomfort of noise and vibration: effects of stimulus duration. Ergonomics, 57, 1244–1255. 10.1080/00140139.2014.914580.

[40] Marfia, G; Guarnaccia, L; Navone, SE; *et. al*. Microgravity and the intervertebral disc: The impact of space conditions on the biomechanics of the spine. Front Physiol. 2023; 14: 1124991. doi: 10.3389/fphys.2023.1124991

[41] Waongenngarm, P., Beek, A., Akkarakittichoke, N., & Janwantanakul, P. (2020). Perceived musculoskeletal discomfort and its association with postural shifts during 4-h prolonged sitting in office workers.. Applied ergonomics, 89, 103225. 10.1016/j.apergo.2020.103225.

[42] Öztürk, N., Öter, E., Abacıgil, F., & Ersungur, E. (2023). Effect of an online posture exercise program during the COVID-19 pandemic on students’ musculoskeletal pain and quality of life. Journal of Back and Musculoskeletal Rehabilitation. 10.3233/bmr-230279.

[43] Johnson, S. (1993). Ergonomic hand tool design. Hand clinics, 9 2, 299–311.

[44] Kalicinski, M; Steinberg, F; Dalecki M, Bock, O. Gaze Behavior While Operating a Complex Instrument Control Task Aerosp Med Hum Perform. 2016 Jul;87(7):646–51. doi: 10.3357/AMHP.4542.2016.

[45] Soto, E; Vega R. Use of galvanic vestibular stimulation device as a countermeasure for microgravity effects in spaceflight. Front. Space Technol., 2024

[46] Oluwafemi FA, Abdelbaki R, Lai JC, Mora-Almanza JG, Afolayan EM. A review of astronaut mental health in manned missions: Potential interventions for cognitive and mental health challenges. Life Sci Space Res (Amst). 2021.

[47] Saniotis, Arthur; G. De la Torre, Gabriel; Galassi, Francesco; Henneberg, Maciej; Mohammadi, Kazhaleh. Mindfulness Meditation and Spaceflight: A Potential Adjunct Therapy for Astronauts. The Mind 2003 3. 10.61936/themind/202312222.

[48] Trindade, Jefferson da Silva; ROQUE, Jessica da Silva Santos. Animais peçonhentos na area rural, 2024. Trabalho de conclusão de curso (Curso Técnico em Segurança do Trabalho) - ETEC Darcy Pereira de Moraes, Itapetininga, 2024.

[49] Alvear, A., Disdier, S., Cruz, R., & Goenaga, M. (2017). Virtual Reality Therapy Implementation for Zoophobia., 448. 10.18687/LACCEI2017.1.1.448.

[50] Lima, Raullyan Borja et al. Espécies vegetais usadas como repelentes e inseticidas no estado do Amapá, BR. Revista Brasileira de Agroecologia, v. 14, n. 3, p. 14–14, 2019.

[51] Agência Nacional de Vigilância Sanitária. (2023). Orientações para os consumidores de saneantes. Recuperado de https://cvs.saude.sp.gov.br/zip/cartilha_de_orientacao_para_os_consumidores.pdf

[52] Kramer, L., & Ciota, A. (2015). Dissecting vectorial capacity for mosquito-borne viruses.. Current opinion in virology, 15, 112–8. 10.1016/j.coviro.2015.10.003.

[53] Wang, L., & Anderson, D. (2019). Viruses in bats and potential spillover to animals and humans. Current Opinion in Virology, 34, 79–89. 10.1016/j.coviro.2018.12.007.

[54] Tavares, Aluska Vieira et al. Epidemiology of the injury with venomous animals in the state of Rio Grande do Norte, Northeast of Brazil. Ciencia & saude coletiva, v. 25, p. 1967–1978, 2020.

[55] Savu, A., Schoenbrunner, A., Politi, R., & Janis, J. (2021). Practical Review of the Management of Animal Bites. Plastic and Reconstructive Surgery Global Open, 9. 10.1097/GOX.0000000000003778.

[56] Instituto Nacional Do Semiárido - Insa. (2013). Cactos do semiárido do Brasil: Guia ilustrado (1^ª^ ed.). Instituto Nacional do Semiárido.

[57] Roberts, D. R. (2012). Extravehicular activity safety manual. NASA Technical Reports.

[58] Costa, A. F. (2018). Urticaceae do Brasil: Taxonomia, distribuição e conservação. Revista Brasileira de Botânica, 41(2), 345-357.

[59] BRITO, Bruno Menezes Nogueira. Correlação entre variáveis meteorológicas e focos ativos para o Rio Grande do Norte. 2021.

[60] Mahaffy, P. R., Webster, C. R., Atreya, S. K., Franz, H., Wong, M., Conrad, P. G., & Team, M. S. L. (2013). Abundance and isotopic composition of gases in the Martian atmosphere from the Curiosity rover. Science, 341(6143), 263-266.

[61] Kieffer, H. H. (2007). Cold jets in the Martian polar caps. Journal of Geophysical Research: Planets, 118(3), 451–470.

[62] Savioli, G; Zanza, C; Longithano, Y. Heat-Related Illness in Emergency and Critical Care: Recommendations for Recognition and Management with Medico-Legal Considerations. Biomedicines. 2022 Oct; 10(10): 2542.

[63] Armstrong, B., & Kricker, A. (2001). The epidemiology of UV induced skin cancer.. Journal of photochemistry and photobiology. B, Biology, 63 1-3, 8–18. 10.1016/S1011-1344(01)00198-1.

[64] Mazzeo, R. S. (2008). Physiological Responses to Exercise at Altitude. Sports Medicine, 38(1), 1–8. doi:10.2165/00007256-200838010-00001

[65] Nagda, N; Hogdson M. Low Relative Humidity and Aircraft Cabin Air Quality. INDOOR AIR 2001. DOI: 10.1034/j.1600-0668.2001.011003200.x

[66] Munster, D; Matar, M; Gokoglu, S; *et. al*. Strategy for Risk Quantification of Spaceflight Crew Health and Performance Using Dynamic Probabilistic Risk Assessment. 53rd International Conference on Environmental Systems. 2024.

[67] Tanaka, K; Nishimura, N; Kawai Y. Adaptation to microgravity, deconditioning, and countermeasures. The Journal of Physiological Sciences volume 67, 271–281 (2017)

[68] Ramachandran, V., Dalal, S., Scheuring, R. A., & Jones, J. A. (2018). Musculoskeletal Injuries in Astronauts: Review of Pre-flight, In-flight, Post-flight, and Extravehicular Activity Injuries. Current Pathobiology Reports, 6(3), 149–158. doi:10.1007/s40139-018-0172-z

[69] Sbissa, Pedro Paulo Mendes et al. Efeito da respiração controlada e da meditação mindfulness sobre a variabilidade da frequência cardíaca. 2014.

